# Effectiveness of Influenza Vaccines in the HIVE household cohort over 8 years: is there evidence of indirect protection?

**DOI:** 10.1101/2021.02.16.21251827

**Authors:** Ryan E. Malosh, Joshua G. Petrie, Amy Callear, Rachel Truscon, Emileigh Johnson, Richard Evans, Latifa Bazzi, Caroline Cheng, Mark S. Thompson, Emily T. Martin, Arnold S. Monto

**Affiliations:** University of Michigan School of Public Health, Department of Epidemiology; US Department of Veterans Affairs; Centers for Disease Control and Prevention, Influenza Division

**Author notes:** Author Contributions:* REM, ETM, and ASM conceptualized the data analysis. REM, ETM, ASM, JGP, MST contributed to the study design. JGP, AC, RE, LB, CC participated in data preparation and cleaning; RT and EJ conducted laboratory assays; all authors participated in the writing and editing of the final manuscript. Disclaimer:* The findings and conclusions in this report are those of the authors and do not necessarily represent the official position of the Centers for Disease Control and Prevention.

## Abstract

**Background:** The evidence that influenza vaccination programs regularly provide protection to unvaccinated individuals (i.e. indirect effects) of a community is lacking. We sought to determine the direct, indirect, and total effects of influenza vaccine in the Household Influenza Vaccine Evaluation (HIVE) cohort.

**Methods:** Using longitudinal data from the HIVE cohort from 2010-11 through 2017-18, we estimated direct, indirect, and total influenza vaccine effectiveness (VE) and the incidence rate ratio of influenza virus infection using adjusted mixed-effect Poisson regression models. Total effectiveness was determined through comparison of vaccinated members of full or partially vaccinated households to unvaccinated individuals in completely unvaccinated households.

**Results:** The pooled, direct VE against any influenza was 30.2% (14.0-43.4). Direct VE was higher for influenza A/H1N1 43.9% (3.9 to 63.5) and B 46.7% (17.2 to 57.5) than A/H3N2 31.7% (10.5 to 47.8); and was higher for young children 42.4% (10.1 to 63.0) than adults 18.6% (−6.3 to 37.7). Influenza incidence was highest in completely unvaccinated households (10.6 per 100 person-seasons) and lower at all other levels of household vaccine coverage. We found little evidence of indirect VE after adjusting for potential confounders. Total VE_T_ was 56.4% (30.1-72.9) in low coverage, 43.2% (19.5-59.9) in moderate coverage, and 33.0% (12.1 to 49.0) in fully vaccinated households.

**Conclusion:** Influenza vaccines may have a benefit above and beyond the direct effect but that effect in this study was small. While there may be exceptions, the goal of global vaccine recommendations should remain focused on provision of documented, direct protection to those vaccinated.

## Introduction

Influenza vaccine is the best way to prevent influenza virus infections and their subsequent complications, including hospitalization and death. Despite a universal recommendation in the United States (US) for annual influenza vaccination of all individuals >6 months old [1], vaccine uptake has been consistently suboptimal, especially in certain groups, at approximately 45% [2]. Seasonal epidemics of influenza continue to cause substantial morbidity and mortality [3,4] and the direct vaccine effectiveness (VE) in those vaccinated varies by season and dominant influenza subtype. On an annual basis, direct VE is most commonly estimated by test negative design studies of medically attended illnesses, which does not account for the entire benefit of influenza vaccination, including protection against mild illness and potential indirect effects through herd immunity.

The classic term herd immunity, formalized in the 1970s and 1980s [5,6], describes a scenario under which population level immunity to an infection is sufficiently high that epidemics become less likely or start to diminish. Vaccinated individuals in the population have immunity from their vaccination and are thus protected from infection (direct VE). Unvaccinated individuals, who are not immune, receive indirect protection, primarily through reduction of the number of infectious individuals at the population level. The indirect effect is, therefore, distinct from the direct effect of the vaccine. The total effect of a vaccine represents the combination of direct and indirect effects [7].

A substantial amount of influenza virus transmission is thought to happen in settings with close and prolonged contact, such as households [8]. This setting is, thus, an ideal place to estimate indirect and total effects. There is some evidence of indirect effects from randomized studies, but these studies are limited by shorter follow up or unique populations. A cluster randomized trial in Hutterite communities in Canada [9] showed substantial indirect effects of inactivated influenza vaccines, surprisingly nearly as high as direct effects when 80% of children were vaccinated [9,10]. A modeling study informed by another randomized trial in Hong Kong estimated that, for influenza B, the indirect protection for household contacts could reach 20% under certain scenarios of household transmission and vaccine coverage [11].

The HIVE Cohort has been used to longitudinally evaluate influenza vaccine effectiveness since 2010. The prospective design of the cohort and active surveillance for respiratory illness presents a unique opportunity to observe both direct and indirect impacts of vaccination. Here we estimate the direct, indirect and total vaccine effectiveness of influenza vaccine in households with children over 8 influenza seasons.

## Methods

### Study Population

We used longitudinal data from the Household Influenza Vaccine Evaluation (HIVE) study collected from 2010-2011 through 2017-2018 influenza seasons. These data include active surveillance for acute respiratory illness (ARI) from 3909 individuals from 911 distinct households, for a total of 9371 person-seasons. Recruitment and retention of participants has been previously described [12]. The HIVE study is approved by the institutional review board at the University of Michigan Medical School.

### Influenza vaccination and household vaccine coverage

Influenza vaccination status was determined by a combination of self-report and documentation from electronic medical records (EMR) and the Michigan Care Improvement Registry (MCIR), as has been previously described [12–15]. We then calculated the proportion of vaccinated household members and the incidence of influenza virus infection each season and longitudinally (i.e. pooled over 8 years). We estimated the crude incidence rate of influenza in households by level of vaccine coverage (i.e. completely unvaccinated, low vaccine coverage [0-50%], moderate vaccine coverage [51-99%], and fully vaccinated). We calculated crude incidence rate, incidence rate ratios and 95% confidence intervals (CI) using the R package *epitools*.

### Influenza infection status

Surveillance for acute respiratory illness was carried out from October through May (2010-11 through 2014-15 seasons) or year-round (2015-16 through 2017-18 seasons), as previously described [12]. Respiratory specimens were tested for influenza by RT-PCR, including A(H3N2), A(H1N1)pdm09 subtypes and B(Yamagata) and B(Victoria) lineages, using protocols from the Centers for Disease Control and Prevention.

### Influenza incidence and direct effect of influenza vaccination

We estimated the direct effects of influenza vaccination by comparing the seasonal incidence rate among vaccinated and unvaccinated individuals. Adjusted incidence rate ratios (aIRR) were estimated from mixed-effect Poisson regression models. VE_D_ was calculated as 1-aIRR*100. Vaccination was modeled as a time varying covariate, with some individuals contributing both vaccinated and unvaccinated person-time. Adjusted models included an offset term to account for person time and included potential confounders (age group, sex, and the Advisory Committee on Immunization Practices (ACIP) defined high-risk conditions [16]).

### Indirect and total vaccine effectiveness: Mini-community design

In this study we considered each household in the HIVE cohort to be a mini-community. The mini-community framework treats the household (or another small unit where contact is sufficient for transmission to occur) as the unit in which indirect and total effects of vaccination are to be estimated [17].

We fitted separate mixed-effects Poisson regression models to estimate VE_I_ and VE_T_, with random effects for household and season. Models were adjusted for potential confounders (age group, sex, and ACIP defined high-risk conditions). To estimate VE_I_ we compared the incidence rate of influenza in unvaccinated individuals in completely unvaccinated households to unvaccinated members of households with higher levels of vaccination (i.e. low [0-50%] or moderate [51-99%] coverage). VE_T_ was estimated by comparing the incidence rate of influenza in vaccinated individuals in completely and partially vaccinated households to unvaccinated individuals in completely unvaccinated households (Figure 1).

**Figure 1.**
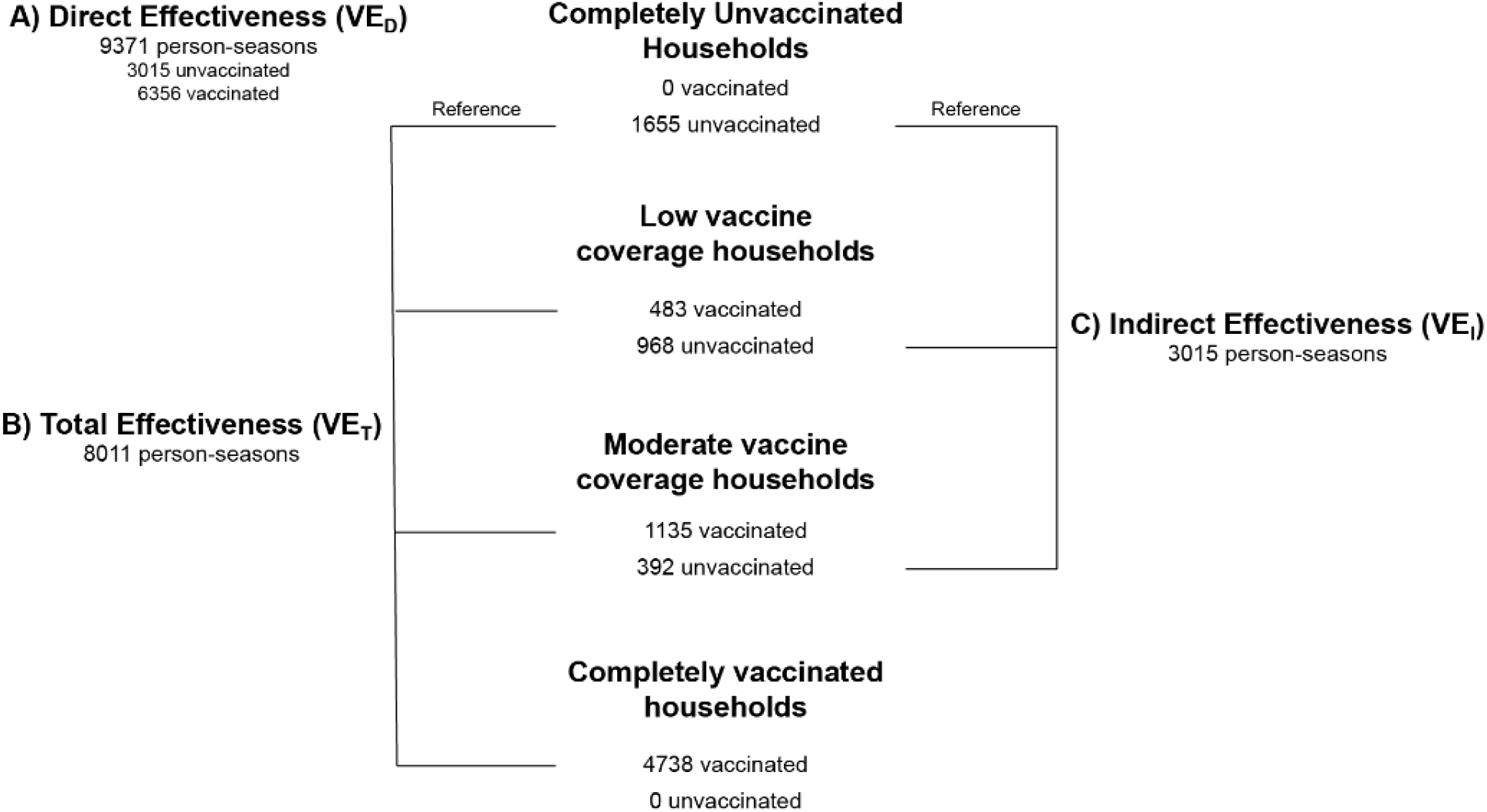
Person-seasons of follow up for comparisons of influenza incidence A) Direct effectiveness (VED) comparing vaccinated and unvaccinated individuals B) Total effectiveness (VET) comparing vaccinated members of moderate and low vaccine coverage households to unvaccinated members of completely unvaccinated households and C) Indirect effectiveness (VEI) comparing unvaccinated members of low and moderate vaccine coverage households to unvaccinated members of completely unvaccinated households, HIVE Study, 2010-2011 through 2017-2018 seasons.

All statistical models were run in R version 4.0.2. Effect estimates were considered statistically significant if 95% confidence interval (CI) did not include the null value.

## Results

We followed 3416 individuals from 799 distinct households, for a total of 9371 person-seasons. Each household was followed for a median of 2 seasons (range 1-8, IQR 1-4). The majority of the observed person-time was in children (58.8%). School-aged children (5-17 years old) in particular, contributed 4,184 (44.7%) person-seasons of follow up. No differences in age distribution were noted by household vaccine coverage. 1753 (48.7%) individuals were female, contributing 4775 (51.0%) person-seasons of follow up. The HIVE cohort is predominantly white, and 16% of participants were considered high risk according to the ACIP definition (Table 1).

**Table 1.**
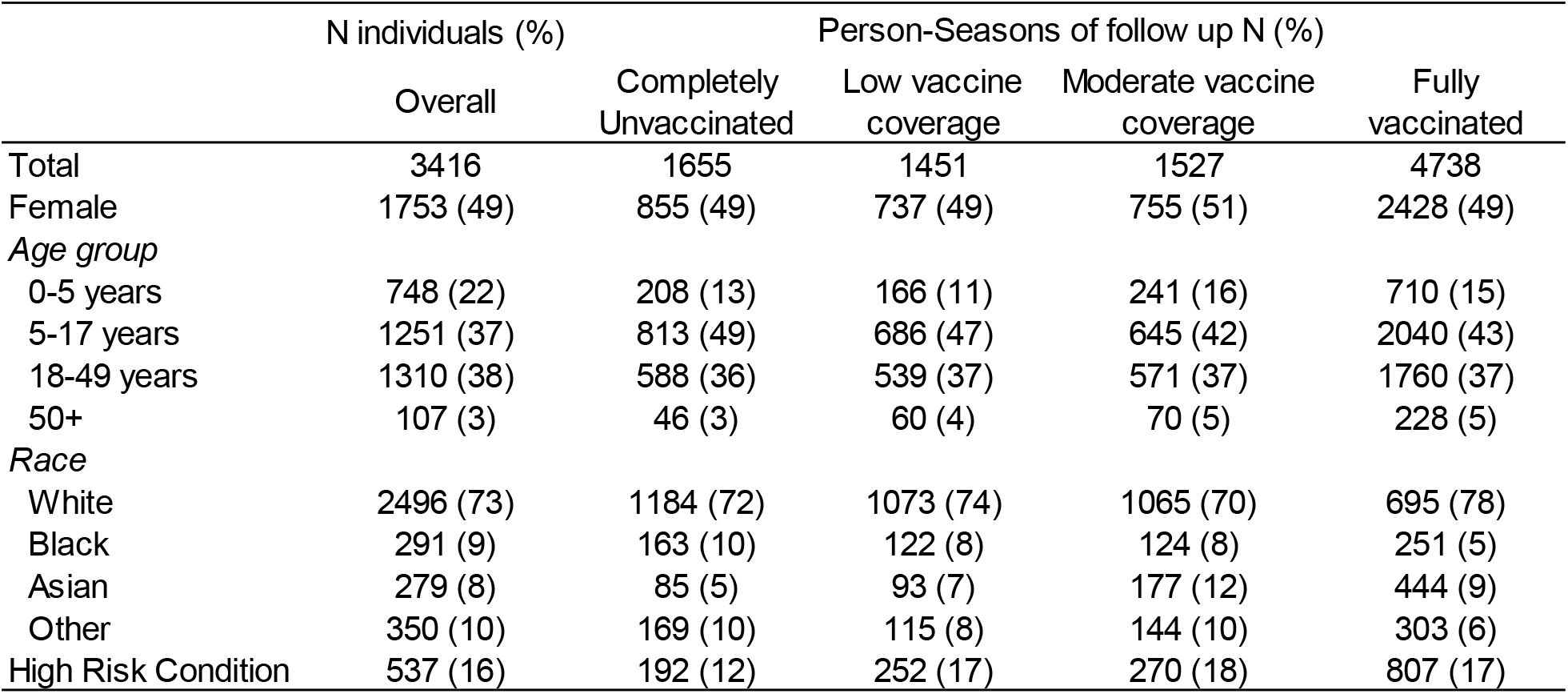
Baseline demographics of HIVE study participants and person-time observed by household vaccine coverage, 2010-2011 through 2017-2018 seasons.

|  | N individuals (%) | Person-Seasons of follow up N (%) |  |  |  |
| --- | --- | --- | --- | --- | --- |
|  | Overall | Completely Unvaccinated | Low vaccine coverage | Moderate vaccine coverage | Fully vaccinated |
| Total | 3416 | 1655 | 1451 | 1527 | 4738 |
| Female | 1753 (49) | 855 (49) | 737 (49) | 755 (51) | 2428 (49) |
| *Age group* |  |  |  |  |  |
| 0-5 years | 748 (22) | 208 (13) | 166 (11) | 241 (16) | 710 (15) |
| 5-17 years | 1251 (37) | 813 (49) | 686 (47) | 645 (42) | 2040 (43) |
| 18-49 years | 1310 (38) | 588 (36) | 539 (37) | 571 (37) | 1760 (37) |
| 50+ | 107 (3) | 46 (3) | 60 (4) | 70 (5) | 228 (5) |
| *Race* |  |  |  |  |  |
| White | 2496 (73) | 1184 (72) | 1073 (74) | 1065 (70) | 695 (78) |
| Black | 291 (9) | 163 (10) | 122 (8) | 124 (8) | 251 (5) |
| Asian | 279 (8) | 85 (5) | 93 (7) | 177 (12) | 444 (9) |
| Other | 350 (10) | 169 (10) | 115 (8) | 144 (10) | 303 (6) |
| High Risk Condition | 537 (16) | 192 (12) | 252 (17) | 270 (18) | 807 (17) |

### Influenza vaccine coverage

Approximately 65% of the HIVE cohort is vaccinated against influenza each year. In total, 6356 (68%) person-seasons among vaccinated individuals over 8 seasons of follow-up were included in this analysis. At the household level, 52% households were completely vaccinated each year, on average (Figure 2). The proportion of households that were completely vaccinated ranged from 47% in 2010-2011 to 60% in 2017-2018. The percentage of households that were completely unvaccinated ranged from a high of 21% in 2010-2011 to a low of 11% in 2017-2018. On average, 17% of households were completely unvaccinated and 31% of households were partially vaccinated each season. There was little variability in vaccine coverage within households among children, pre-school aged children, or school-aged children. In most households, either all children were vaccinated, or none were (Figure S1).

**Figure 2.**
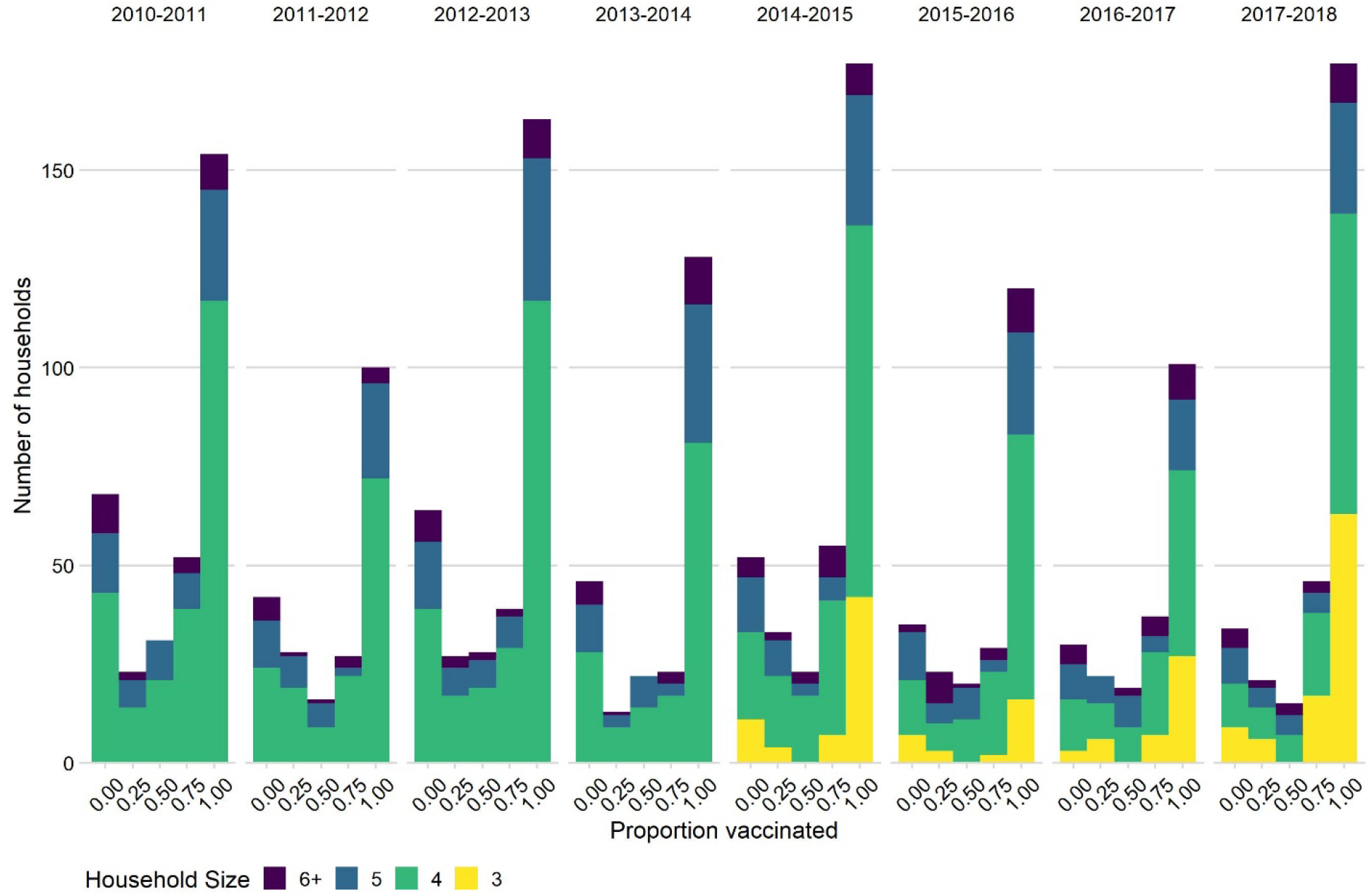
Distribution of household vaccine coverage by household size and season, 2010-2011 through 2017-2018 seasons.

### Influenza incidence and direct effectiveness

The incidence rate of influenza overall was 8.1 per 100 person-seasons (95% CI 7.5-8.7; Table 2). Over the 8 seasons of follow up incidence was highest for influenza A/H3N2 infections (4.6 per 100 person-seasons 95% CI 4.2-5.1), followed by influenza B (2.3 per 100 person-seasons 95% CI 2.0-2.6) and influenza A/H1N1 (1.1 per 100 person-seasons 95% CI 0.9-1.4). Incidence rates for any influenza infection were higher in both pre-school (0-4 years; 10.7 per 100 person-seasons 95% CI 9.0-12.6) and school-aged children (5-17 years; 9.1 per 100 person-seasons 95% CI 8.2-10.1), and lower among adults (≥18 years; 6.1 per 100 person-seasons, 95% CI 5.4-6.9). Seasonal incidence of any influenza virus infection (Table 2) was higher among unvaccinated individuals (10.0 per 100 person-seasons 95% CI 8.9-11.2) than vaccinated individuals (7.2 per 100 person-seasons 95% CI 6.6-7.9). For influenza B, age stratified incidence rates (Table S1) showed substantially higher incidence in pre-school (3.1 per 100 person-seasons) and school-aged children (2.9 per 100 person-seasons) was substantially higher than among adults (1.4 per 100 person-seasons).

**Table 2.** Seasonal incidence rate (95% CI) of influenza infection and direct vaccine effectiveness (VE_D_) overall and by stratified by type/subtype and age group from the HIVE study, pooled over 8 seasons (2010-2011 through 2017-2018 seasons)

|  | Influenza infections | Person-seasons | Seasonal Incidence Rate per 100 person-seasons (95% CI) | Crude VE<sub>D</sub><sup>1</sup> | Unadjusted VE<sub>D</sub> with random effects<sup>2</sup> | Adjusted VE<sub>D</sub> with random effects<sup>3</sup> |
| --- | --- | --- | --- | --- | --- | --- |
| *Any influenza* | 760 | 9371 | 8.1 (7.5-8.7) |  |  |  |
| Unvaccinated | 300 | 3007 | 10.0 (8.9-11.2) | Ref | Ref | Ref |
| Vaccinated | 460 | 6364 | 7.2 (6.6-7.9) | 27.5 (16.2-37.4) | 32.6 (19.8-43.7) | 30.2 (14.0-43.4) |
| **Influenza Type/Subtype<sup>4</sup>** |  |  |  |  |  |  |
| *Influenza A* | 546 | 9371 | 5.8 (5.3-6.3) |  |  |  |
| Unvaccinated | 209 | 3007 | 7.0 (6.0-8.0) | Ref | Ref | Ref |
| Vaccinated | 337 | 6364 | 5.3 (4.7-5.9) | 23.8 (9.5-35.9) | 28.5 (13.0-41.2) | 29.0 (10.0-44.0) |
| *Influenza A/H3N2* | 431 | 9371 | 4.6 (4.2-5.1) |  |  |  |
| Unvaccinated | 161 | 3007 | 5.4 (4.6-6.2) | Ref | Ref | Ref |
| Vaccinated | 270 | 6364 | 4.2 (3.8-4.8) | 20.8 (3.7-34.8) | 27.1 (9.9-41.1) | 31.7 (10.5-47.8) |
| *Influenza A/H1N1* | 107 | 9371 | 1.1 (0.9-1.4) |  |  |  |
| Unvaccinated | 48 | 3007 | 1.6 (1.2-2.1) | Ref | Ref | Ref |
| Vaccinated | 59 | 6364 | 0.9 (0.7-1.2) | 41.9 (15.0-60.3) | 38.7 (0.4-62.3) | 40.7 (3.9-63.5) |
| *Influenza B* | 216 | 9371 | 2.3 (2.0-2.6) |  |  |  |
| Unvaccinated | 91 | 3007 | 3.0 (2.4-3.7) | Ref | Ref | Ref |
| Vaccinated | 125 | 6364 | 2.0 (1.6-2.3) | 35.1 (15.0-50.5) | 37.9 (13.8-55.2) | 46.7 (17.2-57.5) |
| **Age Group** |  |  |  |  |  |  |
| *0-4 years* | 142 | 1325 | 10.7 (9.0-12.6) |  |  |  |
| Unvaccinated | 55 | 325 | 16.9 (12.7-22.0) | Ref | Ref | Ref |
| Vaccinated | 87 | 1000 | 8.7 (7.0-10.7) | 48.6 (28.1-63.3) | 49.7 (26.3-65.7) | 42.4 (10.1-63.0) |
| *5-17 years* | 382 | 4184 | 9.1 (8.2-10.1) |  |  |  |
| Unvaccinated | 163 | 1433 | 11.4 (9.7-13.3) | Ref | Ref | Ref |
| Vaccinated | 219 | 2751 | 8.0 (6.9-9.1) | 30.0 (14.3-42.9) | 33.3 (16.3-46.8) | 28.7 (10.4-43.3) |
| *18+ years* | 236 | 3862 | 6.1 (5.4-6.9) |  |  |  |
| Unvaccinated | 92 | 1257 | 7.3 (5.9-9.0) | Ref | Ref | Ref |
| Vaccinated | 144 | 2605 | 5.5 (4.7-6.5) | 24.5 (1.9-41.9) | 26.5 (4.5-43.4) | 18.6 (-6.3-37.7) |

<sup>1</sup> Direct vaccine effect (VED) calculated as 100\*(1-IRR)
<sup>2</sup> Mixed-effects Poisson regression models with random effects for household and season
<sup>3</sup> Mixed-effects Poisson regression models adjusted for age group, sex, calendar time and high risk condition
<sup>4</sup> Two influenza illnesses were influenza A/influenza B coinfections. These illnesses are considered as one infection for incidence rate calculations of any influenza and separately for type specific incidence rate estimates.

The overall adjusted VE_D_ was 30.2% (95% CI 14.0-43.4) with limited variation by type 40.7% (95% CI 3.9-63.5); 31.7% (95% CI 10.5 to 47.8); 46.7% (95% CI 17.2 to 57.5) for A/H1N1, A/H3N2, and influenza B, respectively). In age-group stratified models, we observed higher levels of protection for pre-school (0-4 years; VE_D_ 42.4%, 95% CI 10.1 to 63.0) and school-aged children (5-17 years; VE_D_ 28.7%, 95% CI 10.4 to 43.3) than we did for adults (≥ 18 years; VE_D_ 18.6, 95% CI −6.3 to 37.7).

### Indirect effectiveness of influenza vaccines

Influenza incidence was generally highest among individuals in completely unvaccinated households (Figure 3). Overall, there appears to be a decline in influenza incidence with increasing proportion of household vaccinated. However, in some seasons, such as 2014-15, when the estimates of VE_D_ in this cohort were essentially zero, the incidence rate was higher than in other seasons and no decline was observed with increasing household vaccine coverage.

**Figure 3.**
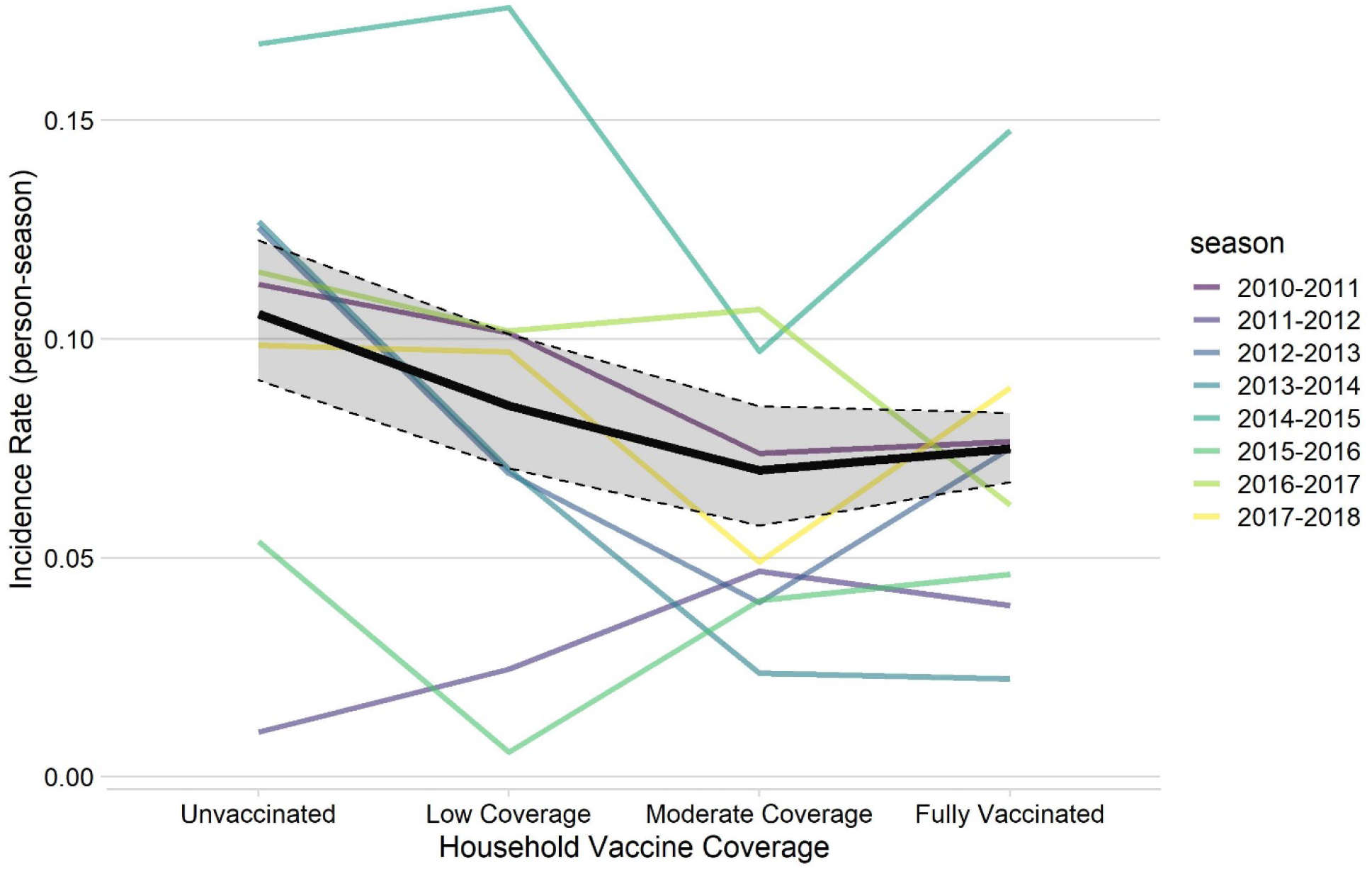
Incidence of influenza infection by household vaccine coverage and season, HIVE Study 2010-2011 through 2017-2018 seasons. Solid lines represent incidence rate estimates, dashed lines represent exact Poisson 95% confidence intervals.

For indirect effect estimates we included a total of 3015 person-seasons of observation in unvaccinated individuals (Figure 1). We observed a lower incidence of influenza among unvaccinated individuals in moderately vaccinated households (8.9 infections per 100 person-seasons [95% CI 6.2-12.4]) compared to those in completely unvaccinated households (10.6 infections per 100 person-seasons [95% CI 9.1-12.3]). Point estimates for crude indirect vaccine effectiveness (VE_i_) comparing unvaccinated individuals in moderate vaccine coverage households to those in completely unvaccinated households suggest low levels of protection but were not statistically different from zero (VE_I_ 15.6 95%CI −21.4 to 41.3). The observed crude indirect VE in low vaccine coverage households was 2.4% (95% CI −24.9 to 24.6; Table 3).

**Table 3.** Seasonal incidence rate (95% CI) of any influenza infection and indirect vaccine effectiveness (VE_I_) in the HIVE study population, pooled over 8 seasons (2010-2011 through 2017-2018 seasons)

|  | Influenza infections | Person-seasons | Seasonal Incidence Rate per 100 person-seasons (95% CI) | Crude VE<sub>I</sub><sup>1</sup> | Unadjusted VE<sub>I</sub> with random effects<sup>2</sup> | Adjusted VE<sub>I</sub> with random effects<sup>3</sup> |
| --- | --- | --- | --- | --- | --- | --- |
| Completely Unvaccinated | 175 | 1655 | 10.6 (9.1-12.3) | Ref | Ref | Ref |
| Low vaccine coverage | 100 | 968 | 10.3 (8.4-12.6) | 2.4 (-24.9 to 24.6) | 9.6 (-21.1 to 32.6) | 8.2 (-20.6 to 30.1) |
| Moderate vaccine coverage | 35 | 392 | 8.9 (6.2-12.4) | 15.6 (-21.4 to 41.3) | 16.9 (-25.2 to 44.9) | 4.6 (-41.2 to 35.6) |

<sup>1</sup> Indirect vaccine effect (VE<sub>I</sub>) calculated as 100\*(1-IRR)
<sup>2</sup> Mixed-effects Poisson regression models with random effects for household and season
<sup>3</sup> Mixed-effects Poisson regression models adjusted for age group, sex, calendar time and high risk condition

In both unadjusted and adjusted models there was no significant reduction in influenza incidence comparing unvaccinated individuals in low or moderately vaccinated households to those in completely unvaccinated households (Table 3). Age-group stratified models (Table S2) demonstrate that school-aged children had lower indirect effect estimates in low (−3.4% 95% CI −52.9 to 30.0) and in moderate (−69.8% 95% CI −216.9 to 9.0) coverage households than either pre-school aged children (low VE_I_ 11.7% 95% CI −63.8 to 52.4 moderate VE_I_ 10.2% 95% CI − 195.3 to 72.7) or adults (low VE_I_ 15.3% 95% CI −38.3 to 48.2; moderate VE_I_ 21.1 95% CI −37.6 to 54.8), suggesting that school-aged children were not as well protected by the vaccination status of their close contacts. Age-stratified models had small sample sizes, however, and the confidence intervals for these VE_I_ estimates were broad and overlapping. We also estimated VE_I_ by influenza type/subtype. We found that the indirect VE was lowest for influenza A/H3N2 (low VE_I_ −32.2% 95% CI −94.4 to 10.1); moderate VE_I_ −19.7% 95% CI −111.4 to 32.2) and highest for influenza A/H1N1 low VE_I_ 56.3% 95% CI −9.2 to 82.5; moderate VE_I_ 24.9% 95% CI - 122.9 to 74.7), matching the VE_D_ estimates (Table S3).

### Total household effect of influenza vaccines

The total effect of influenza vaccines compares vaccinated individuals in households with varying levels of vaccine coverage to unvaccinated individuals in completely unvaccinated households. To estimate total effect of influenza vaccine (VE_T_) we included 8011 person-seasons of follow up in the analytical subset. The crude incidence rate was again highest among individuals in completely unvaccinated households (10.6 per 100 person-seasons 95% CI 9.1-12.3) (Table 4). Among vaccinated individuals in low (5.5 per 100 person-seasons 95% CI 3.6-8.1), moderate (6.8 per 100 person-seasons 95% CI 5.3-8.5), and fully vaccinated (7.9 per 100 person seasons 95% CI 7.1-8.7) households the incidence rate was lower. Notably, there is substantial overlap in the confidence intervals of the incidence rate estimates among vaccinated individuals in households with varying levels of vaccine coverage.

**Table 4.**
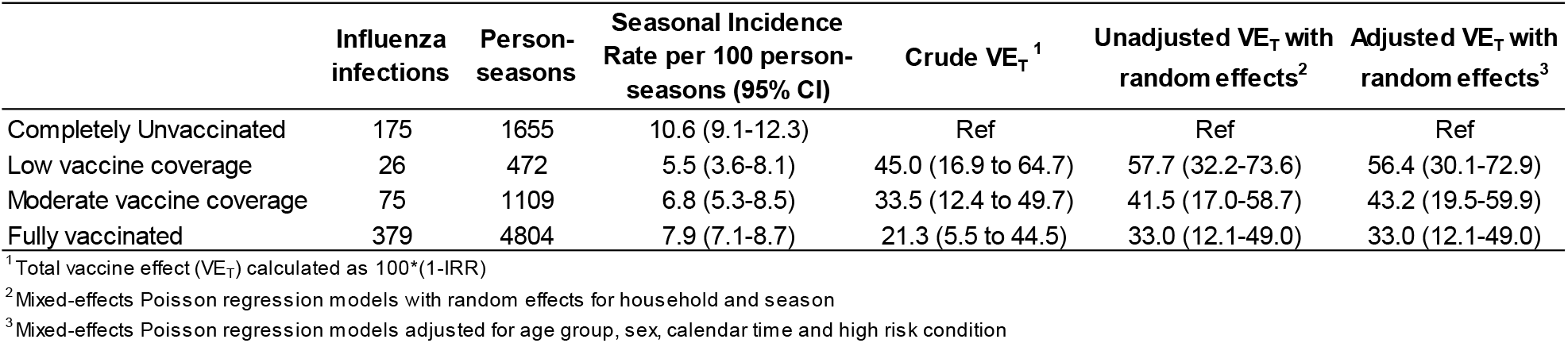
Seasonal incidence rate (95% CI) of any influenza infection and total vaccine effectiveness (VE_T_) in the HIVE study population, pooled over 8 seasons (2010-2011 through 2017-2018 seasons)

We found a significant total effect of influenza vaccines. In low and moderate vaccine coverage households, the VE_T_ was 56.4% (95% CI 30.1 to 72.9) and 43.2% (95% CI 19.5 to 59.9), respectively, after adjusting for potential confounders. For individuals in fully vaccinated households the VE_T_ was also significant, VE_T_ 33% (95% CI 12.1 to 49.0) but this estimate was similar to the overall direct VE.

## Discussion

In this prospective longitudinal study over 8 years, we were able to demonstrate the nearly consistent moderate, direct protection against symptomatic influenza infection by vaccination. This protection has been demonstrated previously, but our findings include mild and moderate illnesses often missed by studies of medically-attended acute respiratory illness (MAARI). This was a result of our use of active surveillance and broad case definition and was particularly of interest for type B infections, which are more difficult to study with the MAARI design. With the exception of most recent seasons, influenza B has typically circulated following an initial influenza A season and often lasts into the spring months, requiring longer duration of annual surveillance to measure. Type B influenza also is more likely to produce illnesses requiring medical attention in children than in younger adults [18], limiting the ability to identify the range of protection. In this study, direct influenza VE was highest for influenza B and for A/H1N1. The low estimates for A/H3N2 reflect global patterns in vaccine effectiveness during this time period and are particularly driven by the 2014-2015 season, when incidence was high, vaccine strain (A/Texas/50/2012) was considered a mismatch with predominant circulating viruses (genetic group 3c.2a), and vaccine effectiveness was near zero [15,19,20].

The household-based design enabled further evaluation of both direct and indirect protection, and in these analyses, we demonstrated significant total effectiveness of vaccination at the household level. However, this effect was statistically similar across households that varied in the extent of vaccination coverage. Indirect protection of unvaccinated people that live in a highly-vaccinated household was not evident. We were able to demonstrate a total effect of vaccine household vaccine coverage that was larger than the direct VE in low and moderate vaccine coverage households. Interestingly, we observed a trend of lower total VE point estimates with increasing household coverage. In fully vaccinated households, for example, total VE (33%) was similar in magnitude to the direct VE (30%), which may suggest that protection in these households is primarily driven by the direct vaccine effects rather than a combination of direct and indirect effects. It is also possible that individuals in fully vaccinated households are more health-conscious and are more likely to report illnesses meeting our case definition and are thus more likely to have influenza detected.

Studies of indirect protection of influenza vaccines have, in general, been limited to ecologic studies [21–24], modeling studies, or relatively small studies with non-specific outcomes (e.g. febrile respiratory illness) [25,26]. Few have been conducted in large populations in natural communities or in communities primarily vaccinated with inactivated influenza vaccine instead of live attenuated influenza vaccine. Previous individual-based studies have demonstrated indirect effects of influenza vaccine, in some cases as large as the direct VE [9,10]. In these studies, the magnitude of the indirect effects has varied based on the predominant circulating viruses. Thus, we explored indirect protection by influenza subtype (Table S3). In particular, we expected lower indirect protection against influenza A/H3N2 viruses for the same reason that we expected lower direct VE. Our findings suggest that indirect protection, if present, may be higher for influenza A/H1N1 but the small numbers in these stratified analyses limit our ability to draw conclusions.

We also wanted to explore if indirect effects varied by age-group. In age-group stratified models, we did see potential effect modification of the indirect protective effect. In particular, the lack of an observed indirect effect seems to be driven by the fact that school-aged children did not benefit from vaccination of household contacts in the same way that adults and pre-school aged children did (Table S2). School-aged children are known to drive influenza epidemics [27,28], and as a result of their contact patterns [29], they represent the group at highest risk of community-acquired influenza. In addition, a number of recently observed issues affecting estimates of direct influenza vaccine effectiveness (VE) may also influence the indirect effects. Repeated annual vaccination, antigenic drift, mutations induced by growing vaccine viruses in chicken eggs, and waning immunity have all been linked to lower than expected direct VE in recent years [14,15,19,30–34]. These mechanisms require further exploration as a potential explanation for the lack of indirect VE observed in this analysis.

Lack of heterogeneity in vaccine uptake is a challenge for evaluating indirect protection at the household level, as individuals tend to share vaccination habits with others in their household. Most influenza infections in the HIVE study are considered community-acquired [13,14], making identification of indirect protection resulting from reductions in household transmission risk a challenge. A more granular evaluation of indirect protection at the household level requires a situation where household members have similar risk of infection but different access to vaccination based on individual factors that are not shared by all household members. This is the situation that we are presented with given the U.S. prioritization schemes for SARS-CoV-2 vaccine deployment. Although the household cohort was somewhat limited in the ability to evaluate potential indirect protection for unvaccinated members of partially vaccinated households, future findings in the era of COVID-19 may be quite different. Especially, as prioritization schemes will result in varied vaccination timing among household members.

While we were unable to convincingly demonstrate that there was indirect protection beyond the direct effect, we did show that vaccine had a clear role in preventing even mild disease in a cohort of families living in their own homes. The indirect effect might have been clear if the vaccines had higher direct effects. This is agreement with a recent literature reviews and meta-analyses which found inconsistency in demonstration of indirect effects [35,36]. The whole concept of indirect protection or herd immunity has become a focus in our efforts to control Covid-19 outbreaks. It is important to remember that the primary focus with a vaccine should be good direct VE and that indirect protection should be seen as a bonus, but not a necessity.

## Data Availability

The data included in this analysis are available from the corresponding author on reasonable request.

## Acknowledgments

The authors gratefully acknowledge the contribution of the HIVE study households. We also recognize the efforts of current and past HIVE Study staff Anne Kanicledes, Barbara Aaron, and Casey Martens; past study investigators Suzanne Ohmit; and current and past students Hannah Segaloff, Peter DeJonge, Kat Miller, and Khalil Chedid.

## Supplemental Material

### Household vaccine coverage by age group

**Figure S1.**
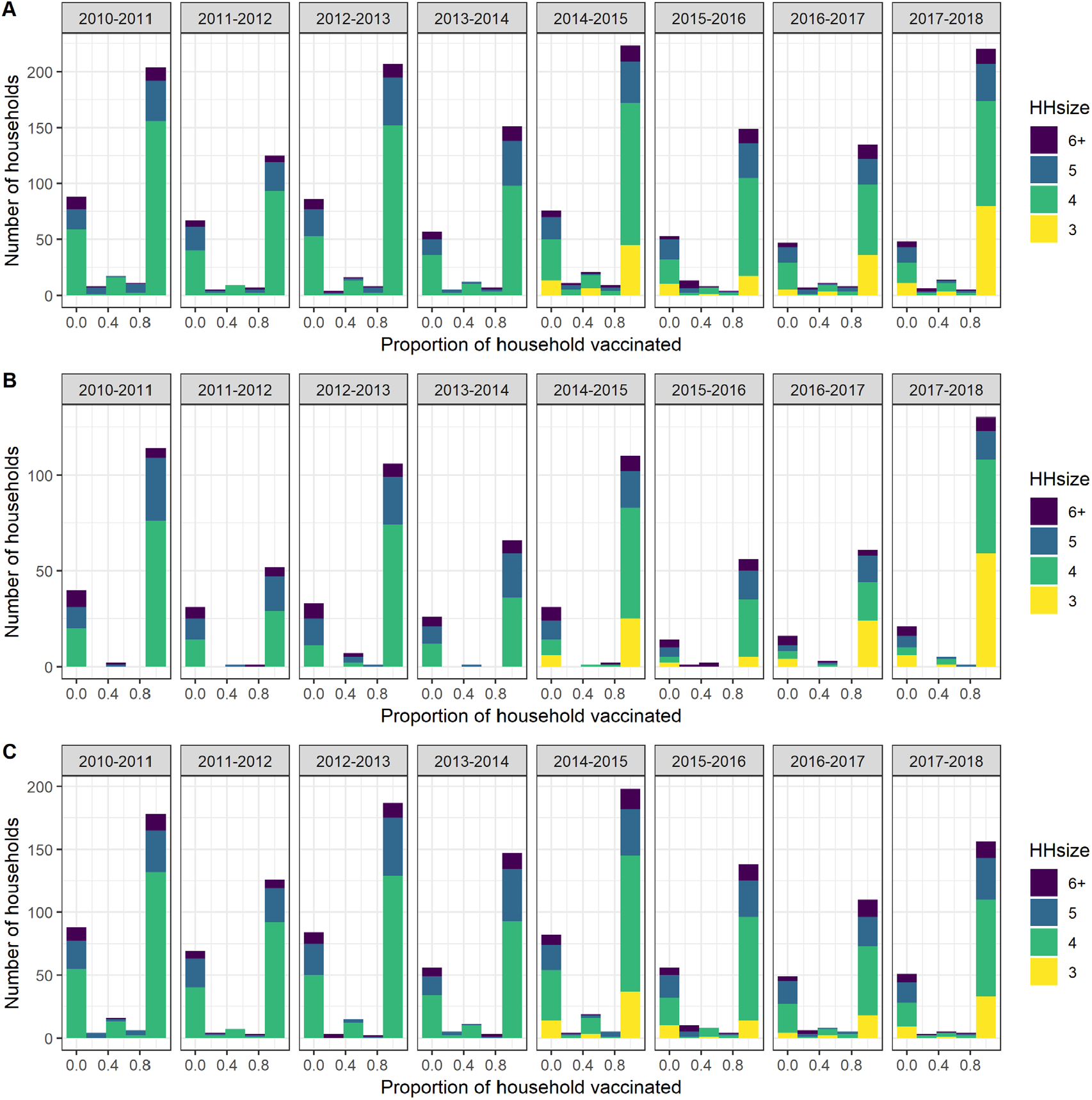
Distribution of household vaccine coverage by age group vaccinated A) All children < 18 years B) Pre-school aged children (i.e. children < 5 years), and C) School-aged children (i.e. children 5-17 years), HIVE Study 2010-2011 through 2017-2018 seasons

### Age group stratified incidence of influenza infection by type

**Table S1.**
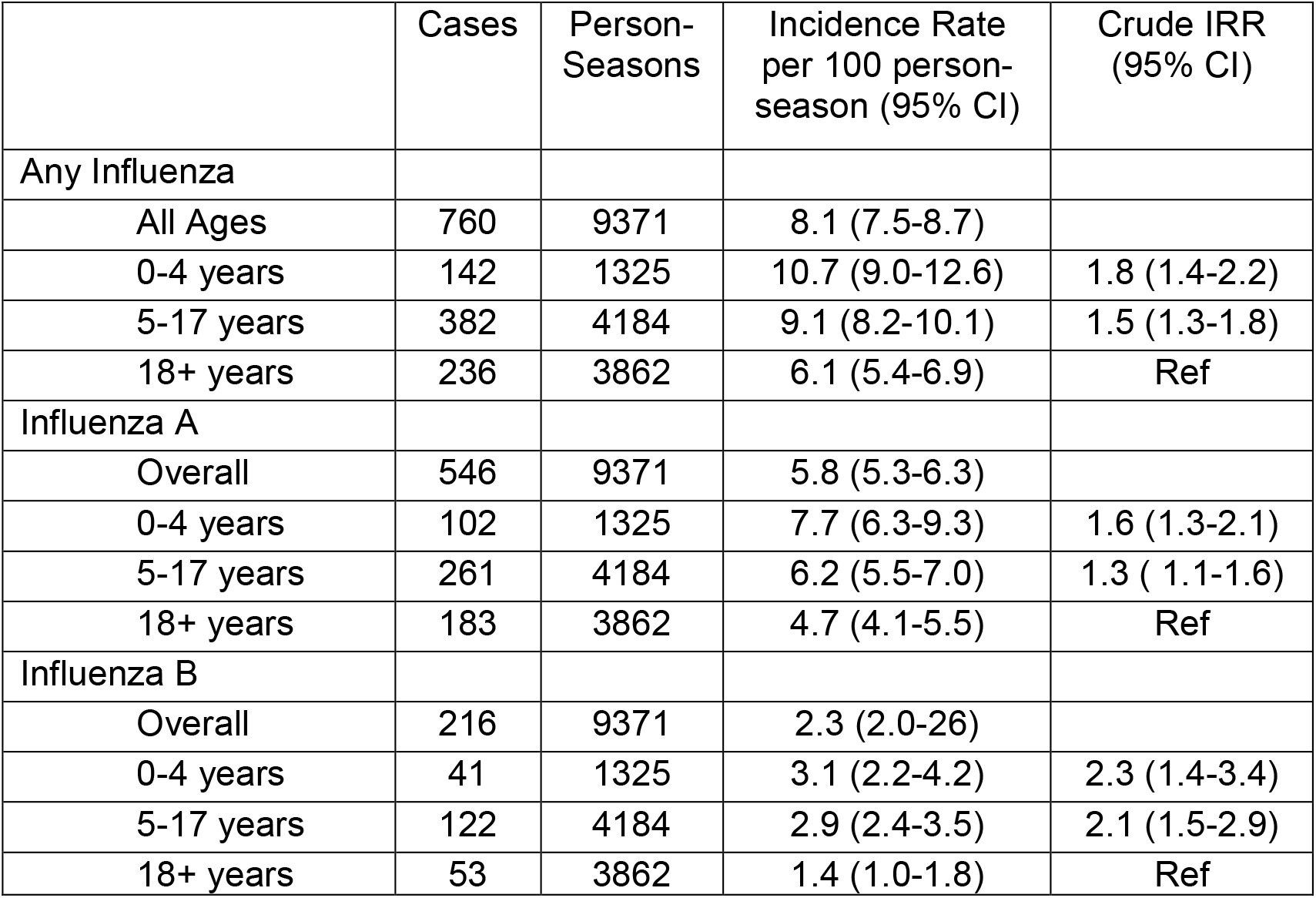
Incidence rate of influenza virus infection by type and age group and crude incidence rate ratios (IRR) for age group, HIVE Study 2010-2011 through 2017-2018 seasons

### Age group stratified models of indirect protection

Index cases of influenza in a household setting are commonly school aged-children. To explore if age group was an effect modifier for indirect protection, we estimated the indirect VE in separate models for individuals 0-4 years old (pre-school aged children), 5-17 years old (school-aged children) and adults (18+ years).

**Table S2.**
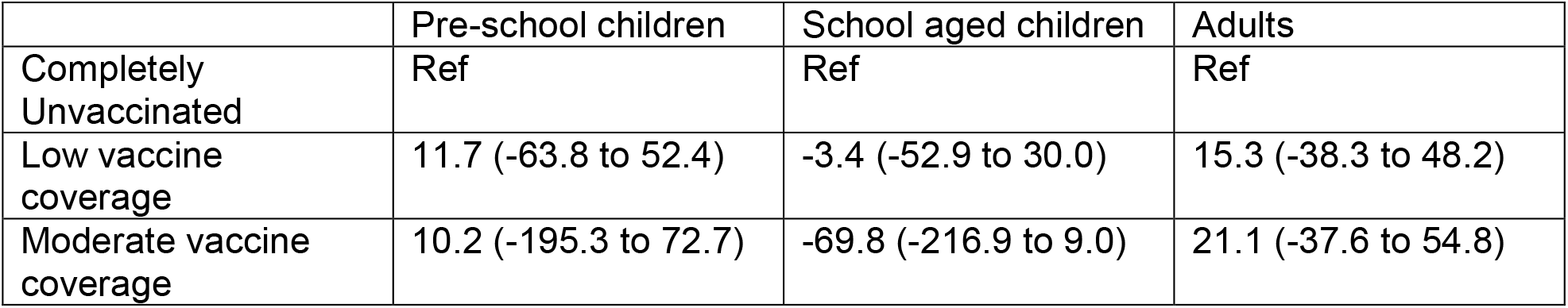
Indirect Vaccine Effect (VE_I_) and 95% CI of household coverage from mixed effect Poisson Models with random effects for individual, household and season, stratified by age group, HIVE Study 2010-2011 through 2017-2018 seasons

### Influenza type/subtype stratified models of indirect protection

**Table S3.** Indirect Vaccine Effectiveness (VE_I_) and 95% CI of household coverage from mixed effect Poisson Models with random effects for individual, household and season, stratified by influenza type/subtype,, HIVE Study 2010-2011 through 2017-2018 seasons

|  | Influenza A/H3N2 | Influenza A/H1N1 | Influenza B |
| --- | --- | --- | --- |
| Completely Unvaccinated | Ref | Ref | Ref |
| Low vaccine coverage | -32.2 (-94.4 to 10.1) | 56.3 (-9.2 to 82.5) | 41.0 (-6.3 to 67.3) |
| Moderate vaccine coverage | -19.7 (-111.4 to 32.2) | 24.9 (-122.9 to 74.7) | 9.7 (-107 to 60.7) |

